# The relationship between postpartum care uptake and postpartum morbidity and their determinants in Morocco: evidence from a national survey

**DOI:** 10.1101/2025.09.08.25334839

**Authors:** Asmaa Habib, Hafiz T.A. Khan, Caroline Lafarge, Rachid Bezad

**Affiliations:** Public health Group, University of West London, Boston Manor Road, Brentford, Middlesex TW8 9GB, United Kingdom; Faculty of Medical Sciences, UM6P Hospitals, Mohammed VI Polytechnic University, Benguerir, 43150, Morocco; Graduate School, University of West London, St Mary’s Road, London W5 5RF, United Kingdom; Faculty of Medicine and Pharmacy, University Mohamed V, Rabat Morocco

**Keywords:** Postpartum care, postpartum morbidity, maternal health, health inequities, Morocco

## Abstract

**Background:** PPC utilisation is essential to prevent maternal mortality and morbidity, particularly in low-and middle-income countries where 95% of maternal deaths occur. In Morocco, PPC remain underused. This study examines factors associated with PPC utilisation and postpartum morbidity (PPM), and their relationship.

**Methods:** A secondary analysis was conducted using a nationally representative dataset of 5,593 women of childbearing age. Univariate and multivariate logistic regression assessed the associations of sociodemographic, environmental and obstetrical determinants with PPC and PPM.

**Results:** 62.6% of women reported early PPC (EPPC), 21.3% used later PPC (LPPC) and 28.3% declared PPM. Facilitators associated with LPPC included education above primary level (AOR=1.34, 95%CI:1.11-1.63), high socioeconomic status (AOR=1.42, 95%CI:1.02-1.98), antenatal care (AOR=1.64, 95%CI:1.08-2.47), caesarean delivery (AOR=2.50, 95%CI:1.89-3.31), and newborn postnatal care (AOR=6.97, 95%CI:5.89-8.25). Absence of doctors during midwives-led delivery reduced LPPC uptake (AOR=0.63, 95% CI:0.48-0.83). Secondary or higher education (AOR=0.71, 95%CI:0.54-0.93) and antenatal care (AOR=0.30, 95%CI:0.14-0.64) reduced PPM risk, while instrumental delivery (AOR=1.24, 95%CI:1.04-1.48) and pregnancy morbidities (AOR=2.10, 95%CI:1.72-2.56) increased it. EPPC lowered PPM risk (AOR=0.65, 95%CI:0.52-0.79), whereas LPPC utilisation was associated with PPM (AOR=1.36, 95%CI:1.08-1.71).

**Conclusion:** LPPC utilisation remains low, and PPM persists, reflecting health inequities. Targeted interventions for disadvantaged women and further qualitative research into behavioural and cultural influences, and women’s and health professionals’ perceptions of PPC are recommended.

## INTRODUCTION

Postpartum care (PPC) utilisation is essential to prevent postpartum maternal mortality and morbidity, particularly in low-and-middle income countries where 95% of maternal death occur (1). The World Health Organization recognizes the importance of using postpartum care (PPC) consultations to screen and raise awareness of symptoms of the most common postpartum morbidities (PPM) and inform women about good postpartum practices. Sepsis (including metritis which is the infection of the uterus during or after labour), haemorrhage, and hypertensive disorders (eclampsia) were the most life-threatening conditions and were responsible for over half of maternal deaths in the world (2,3). More than two thirds of haemorrhage-related deaths occurred within six weeks after childbirth (3). In Morocco, research on PPM started in 2005 (4) but few studies have since investigated this topic. In the latter, the complications reported by women included haemorrhage (79.9%), fever (12.1%), pregnancy-related hypertension (10.6%), mental disorders (10%), genital infections (8%), and breast conditions (5%) (5,6). Moreover, PPC utilisation has remained low and stagnant since 2011 with only 21.8% of women reporting attending PPC consultations (7,8). The literature showed that several factors are associated with PPC uptake including women’s education, socioeconomic status, place of residence, antenatal care visit, health facility based-delivery, caesarean delivery, skilled-birth deliveries, women’s autonomy in decision making (9,10).

The aim of this study was, to determine the scope of PPC uptake in Morocco and understand its barriers and facilitators determinants. The second objective was to assess the relationships between PPC uptake and PPM occurrence in Morocco.

## Methods

### Study design and population

The ethical approval to conduct this study was given by the University of West London, the 10^th^ February 2020 (No.00748). The study was based on secondary data analysis of the women’s health questionnaire (sections 1, 3 and 3) of the *Enquête Nationale sur la Population et la Santé Famililale* - National Survey of Population and Family Health 2018 which is a large-scale cross-sectional survey used in Morocco (8). The questionnaire is constructed from the Multiple Indicator Cluster Surveys (MICS) and the Pan Arab Project for Family Health (PAPFAM) questionnaires and adapted to the Moroccan context. The sampling methods used (i.e. two-stage stratified probability sampling method) ensure the representativeness of the data to the total population of the country.

Eligible participants were women aged between 15 and 49 years, who gave birth to a live baby within five years prior to the survey (from 2013 to 2017) and who were non-single (meaning married, divorced, separated, or widowed) at the time of the survey (supplementary file 1). The fact that only non-single women were recruited did not affected the representativeness of the data because the majority of women who deliver in Morocco are not single (in 2018, 58.5% married, 9.9% widow, 3.4 divorced, 0.6% separated) (8).

### Outcome variables

Three dependent variables were analysed. The first one was ‘early PPC uptake’(EPPC) before discharge from hospital. Analyses related to this dependent variable excluded women who delivered at home or other places. The second variable was ‘later PPC uptake’ (LPPC) within six weeks post-delivery. The third variable was PPM occurrence which included the following physical health complications: acute vaginal haemorrhage, oedema and foot pain, smelly vaginal discharge with fever, pelvic pain with fever, lower back pain with fever, dorsal pain with fever, urinary burning with fever, pain and swelling mammary with fever, and other morbidities that were not defined in the database. All the outcome variables were binary (Yes/No).

### Independent variables

Altogether, 55 predictors (or independent variables) were analysed and classified into four main categories: sociodemographic (eight variables), environmental (four variables), obstetric (27 variables) and ‘other’ (16 variables). In this article, only predictors reported in the literature and those included in the multivariate analyses are presented. Among the obstetric predictors the pregnancy health complications variable considered abnormal swelling of the face, fingers, and feet; vaginal haemorrhage; convulsion not caused by fever; intense and persistent headache; blurry vision; intense pelvic pain; hyperventilation; fever with difficulty standing up; and water break six hours before labour.

### Statistical analysis

Five of the 55 predictors had entries with missing data, which were removed prior to analysis. To describe the population of study, the distributions of dependent and independent variables were assessed. Moreover, univariate analyses were performed to estimate the associations between predictors and EPPC, LPPC and PPM, independently. For predictors with two modalities, Chi-squared tests were carried out, whereas bivariate analyses were chosen to examine the associations with predictors defined by at least three categories. The effect sizes were expressed by crude odds ratios with a 5% significance level.

Secondly, a multivariate regression was conducted independently for each of the three outcome variables, which enabled their inclusion as independent variables in the models. We considered a diagnostic test of multicollinearity (i.e., variance inflation factor) to determine the predictors for inclusion in the logistic regression. Only predictors that did not induce serious multicollinearity (11) and already used in the literature (10,12) were deemed eligible to generate reliable statistical regression models. A hierarchical multiple regression method was used with in model 1, sociodemographic and environmental predictors entered to control for their possible confounding bias. In model 2, obstetric predictors were added to measure their effect on the dependent variables above and beyond that of predictors from model 1. The results were reported as adjusted odds ratios, estimated with a 5% level of statistical significance. The analyses were performed using the Statistical Package for the Social Sciences software IBM SPSS V28 (13).

## RESULTS

### Characteristics of respondents

Altogether, 5,593 women were included in this study (supplementary file 1). Analyses related to EPPC applied to women who delivered in a health facility, accounting for 4,792 women. Participants’ mean age was 31.7 (SD:6.8); most women were married (97.5%), unemployed (90.0%), and without formal education (57.8%) or educated to a primary level (32.8%). Only few (9.4%) reached secondary and higher education (Table 1).

**Table 1.** Distribution of women’s characteristics.

| **Characteristics** | **Frequency**<br>**(n)** | **Percent**<br>**(%)** |
| --- | --- | --- |
| **PPC uptake before discharge from health facility (EPPC)** |  |  |
| Yes | 3000 | 53.6 |
| No | 1792 | 32.0 |
| **PPC uptake within 6 weeks post-delivery (LPPC)** |  |  |
| Yes | 1219 | 21.8 |
| No | 4372 | 78.2 |
| **Postpartum morbidity** |  |  |
| No | 4010 | 71.7 |
| Yes | 1583 | 28.3 |
| **Maternal age** |  |  |
| 15-29 | 2289 | 40.9 |
| 30-39 | 2520 | 45.1 |
| 40-49 | 783 | 14.0 |
| **Maternal education** |  |  |
| No formal education | 3235 | 57.8 |
| Primary education | 1835 | 32.8 |
| Secondary/ higher education | 523 | 9.4 |
| **Partner education level** |  |  |
| Primary | 1851 | 44.0 |
| Preliminary/Moderate | 1133 | 27.0 |
| Secondary/ Higher | 1220 | 29.0 |
| **Maternal employment status** |  |  |
| Unemployed | 5035 | 90.0 |
| Employed | 558 | 10.0 |
| **Socioeconomic status** |  |  |
| Poorest | 1275 | 22.8 |
| Poorer | 1146 | 20.5 |
| Middle | 1167 | 20.9 |
| Richer | 1128 | 20.2 |
| Richest | 876 | 15.7 |
| **Marital status** |  |  |
| Married | 5452 | 97.5 |
| Widow | 41 | 0.7 |
| Divorced | 72 | 1.3 |
| Separated | 28 | 0.5 |
| **Place of residence** |  |  |
| Urban | 3128 | 55.9 |
| Rural | 2464 | 44.1 |
| **Long distance from a health facility preventing LPPC uptake** |  |  |
| Yes | 122 | 2.8 |
| No | 4251 | 97.2 |
| **Antenatal care** |  |  |
| 0 visit | 701 | 12.5 |
| 1 to 3 visit(s) | 1926 | 34.4 |
| 4 visits | 1207 | 21.6 |
| > 4 visits | 1759 | 31.4 |
| **Mode of delivery** |  |  |
| Vaginal delivery | 1835 | 38.2 |
| Vaginal delivery assisted by instruments | 1966 | 40.9 |
| Caesarean delivery | 1008 | 21.0 |
| **Place of delivery** |  |  |
| Home | 770 | 13.8 |
| Public hospital | 3162 | 56.8 |
| Delivery centre / health centre | 760 | 13.7 |
| Private clinic | 811 | 14.6 |
| Private surgery | 62 | 1.1 |
| **Length of stay in the health facility after birth** |  |  |
| < a day (hours) | 461 | 9.6 |
| 1 ≥ days ≤ 7 | 4278 | 89.3 |
| ≥ 1 week | 53 | 1.1 |

### Utilisation of postpartum care and postpartum morbidity prevalence

About 62.6% of women received EPPC during delivery-led hospitalisation, but only 21.3% used LPPC within six weeks post-delivery. LPPC were essentially taken in public health facilities or delivery centres (40.0%), and in private surgeries (30.2%) (Table 1). Conversely, 78.2% of women did not use LPPC. The main reasons included the absence of PPM (70.63%), lack of awareness of PPC importance (15.23%), financial difficulty (7.51%), distance to health facility (2.79%), unavailability of LPPC services (1.71%), and other unspecified reasons.

Overall, 28.3% of women reported at least one PPM, with most reporting only a single symptom (15.7%) (Table 1).

### Factors associated with early postpartum care uptake

The findings regarding EPPC uptake are displayed in Table 2.

**Table 2.**
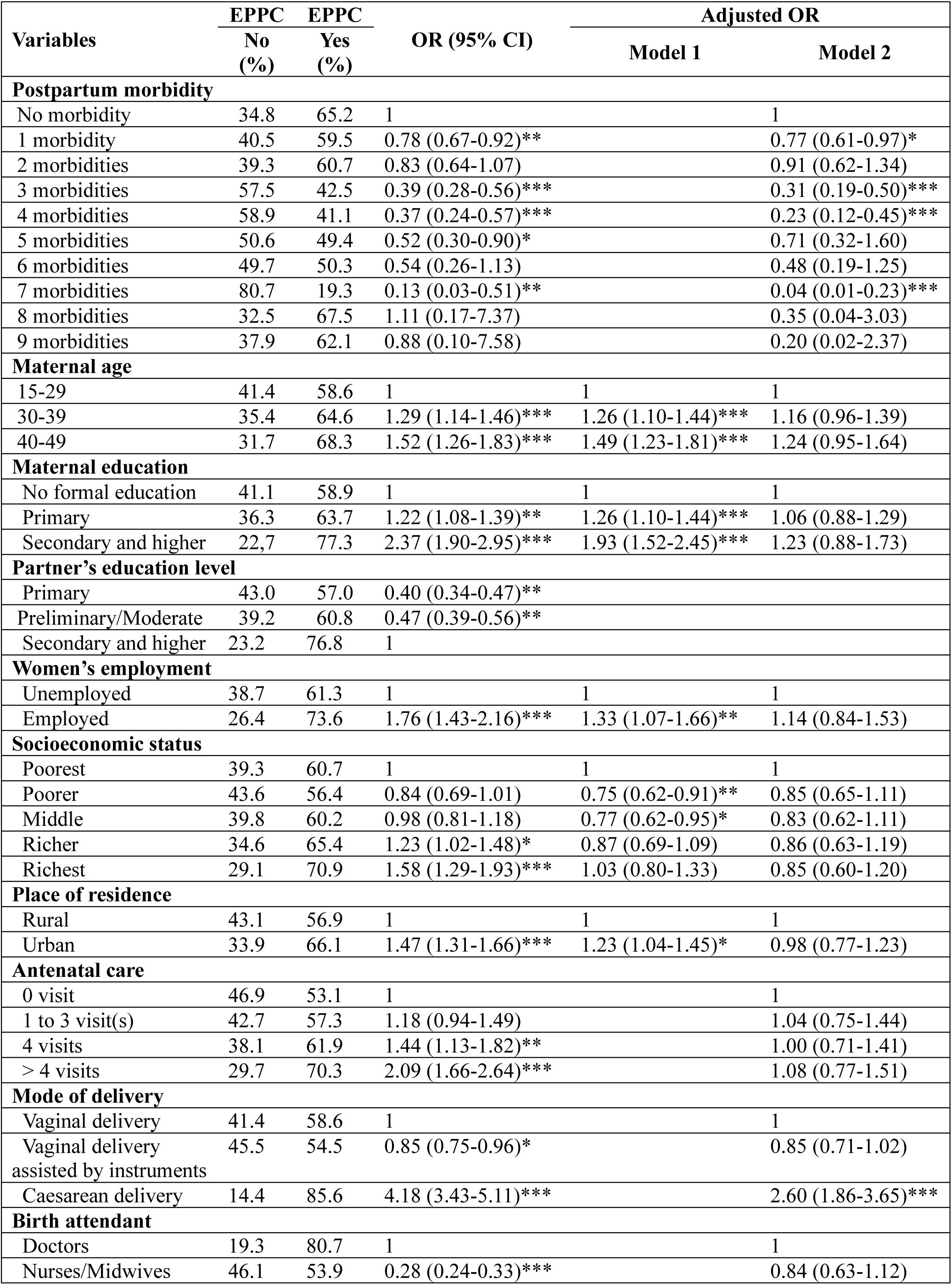
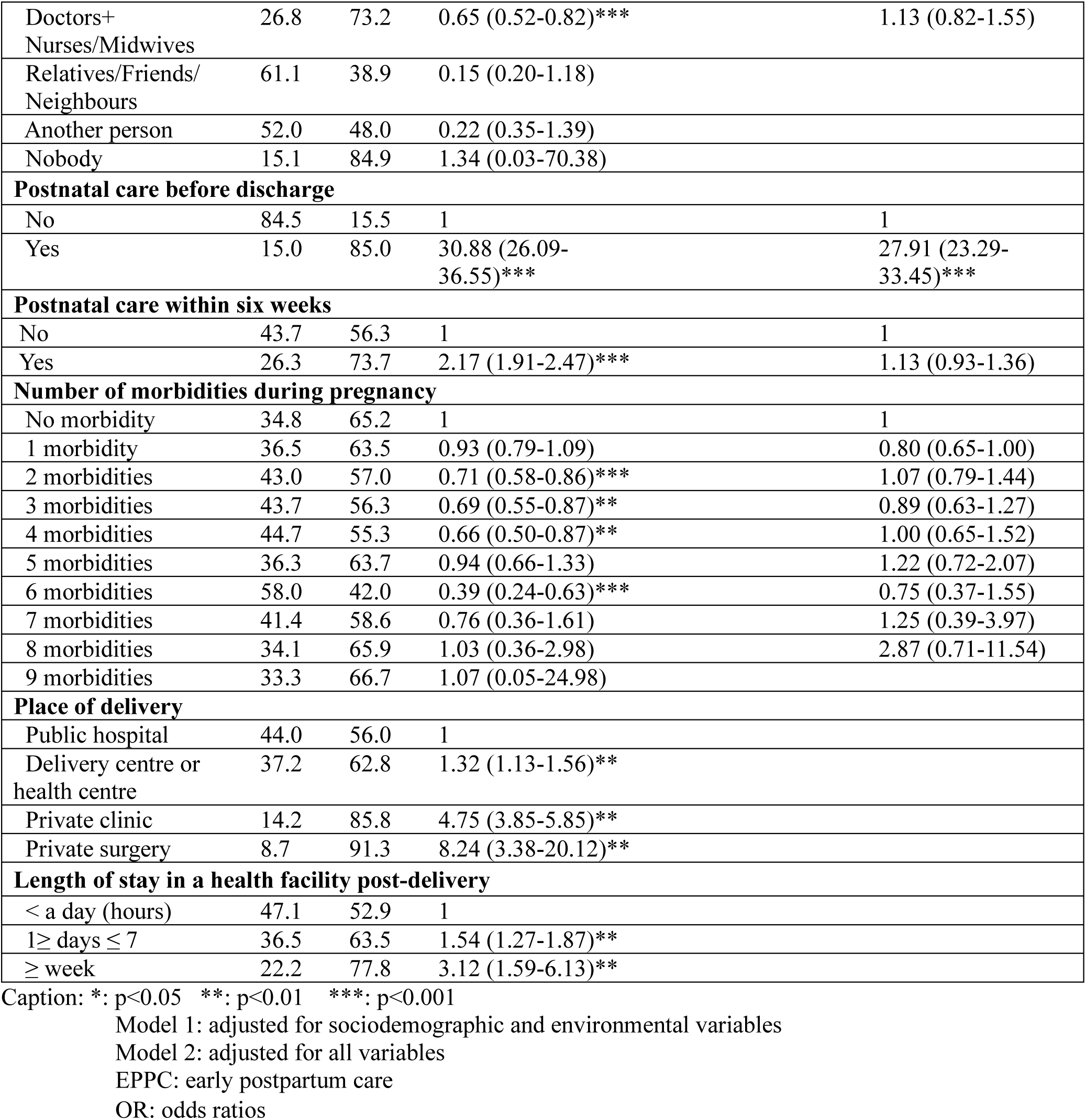
Multilevel logistic regression of factors associated with early postpartum care uptake.

Maternal sociodemographic characteristics associated with EPPC in Morocco were being aged 30-49 years old, having completed formal education (regardless of the level for women and at least of secondary/ higher level for their husbands), being employed, and living in richer or the richest households. However, the logistic regression (Table 2) suggests that when taken into consideration all predictors alongside obstetric factors (model 2), sociodemographic factors were no longer significant predictors of EPPC.

Urban place of residence, attending at least four antenatal care visits and experiencing health complications during pregnancy were additional predictors initially found to facilitate EPPC uptake, but this was not confirmed by the multivariate analysis (Table 2).

In these analyses (Table 2-model 2), delivery by caesarean section was significantly associated with EPPC. Women who delivered by a caesarean section were 2.6 times more likely to receive EPPC compared to their counterparts who had a straight vaginal delivery. Newborn postnatal care (PNC) before discharge was another significant predictor of EPPC uptake as it multiplied by 27.91 times the likelihood of women receiving EPPC for themselves. Thus, the PNC is an opportunity for health professionals to examine both women and babies at the same time.

### Factors associated with later postpartum care uptake

Table 3 presents findings regarding LPPC uptake.

**Table 3.** Multilevel logistic regression of factors associated with later postpartum care uptake in Morocco.

| Variables | LPPC | LPPC | OR (95% CI) | Adjusted OR |  |
| --- | --- | --- | --- | --- | --- |
|  | No (%) | Yes (%) |  | Model 1 | Model 2 |
| Early postpartum care |  |  |  |  |  |
| No | 91.0 | 9.0 | 1 |  | 1 |
| Yes | 68.7 | 31.3 | 4.61 (3.85-5.52)*** |  | 2.68 (2.08-3.45)*** |
| Postpartum morbidity |  |  |  |  |  |
| No morbidity | 79.9 | 20.1 | 1 |  | 1 |
| 1 morbidity | 73.1 | 26.9 | 1.46 (1.23-1.73)*** |  | 1.36 (1.08-1.71)** |
| 2 morbidities | 73.1 | 26.9 | 1.46 (1.12-1.90)** |  | 1.24 (0.86-1.81) |
| 3 morbidities | 76.0 | 24.0 | 1.25 (0.85-1.84) |  | 1.14 (0.64-2.03) |
| 4 morbidities | 75.4 | 24.6 | 1.30 (0.83-2.03) |  | 1.19 (0.59-2.40) |
| 5 morbidities | 78.0 | 22.0 | 1.12 (0.62-2.00) |  | 0.38 (0.16-0.95)* |
| 6 morbidities | 77.9 | 22.1 | 1.12 (0.50-2.50) |  | 0.93 (0.30-2.91) |
| 7 morbidities | 84.0 | 16.0 | 0.76 (0.25-2.30) |  | 0.29 (0.04-2.41) |
| 8 morbidities | 61.5 | 38.5 | 2.48 (0.61-10.13) |  | 3.20 (0.32-31.96) |
| Postpartum pelvic pain with fever |  |  |  |  |  |
| No | 78.6 | 21.4 | 1 |  | 1 |
| Yes | 73.5 | 26.5 | 1.32 (1.05-1.65)** |  | 2.11 (1.35-3.31)** |
| Other postpartum morbidities |  |  |  |  |  |
| No | 78.9 | 21.1 | 1 |  | 1 |
| Yes | 62.4 | 37.6 | 2.25 (1.72-2.95)*** |  | 2.03 (1.38-3.00)*** |
| Maternal age |  |  |  |  |  |
| 15-29 | 81.5 | 18.5 | 1 | 1 | 1 |
| 30-39 | 76.1 | 23.9 | 1.38 (1.20-1.58)*** | 1.42 (1.22-1.67)*** | 1.23 (1.02-1.48)* |
| 40-49 | 75.2 | 24.8 | 1.45 (1.19-1.75)*** | 1.54 (1.23-1.92)*** | 1.17 (0.90-1.52) |
| Maternal education |  |  |  |  |  |
| No formal education | 83.6 | 16.4 | 1 | 1 | 1 |
| Primary | 74.5 | 25.5 | 1.75 (1.52-2.01)*** | 1.71 (1.45-2.01)*** | 1.34 (1.11-1.63)** |
| Secondary and higher | 57.8 | 42.2 | 3.71 (3.05-4.52)*** | 2.99 (2.38-3.75)*** | 1.79 (1.35-2.36)*** |
| Partner's education level |  |  |  |  |  |
| Primary | 82.5 | 17.5 | 1 |  |  |
| Preliminary/Moderate | 78.9 | 21.1 | 1.26 (1.04-1.51)* |  |  |
| Secondary and Higher | 63.1 | 36.9 | 2.75 (2.33-3.25)** |  |  |
| **Women's employment status** |  |  |  |  |  |
| Unemployed | 71.0 | 29.0 | 1 | 1 | 1 |
| Employed | 79.0 | 21.0 | 1.53 (1.26-1.86)*** | 1.01 (0.81-1.26) | 0.81 (0.63-1.06) |
| **Socioeconomic status** |  |  |  |  |  |
| Poorest | 86.2 | 13.8 | 1 | 1 | 1 |
| Poorer | 81.6 | 18.4 | 1.40 (1.13-1.75)** | 1.18 (0.91-1.53) | 1.33 (0.99-1.80) |
| Middle | 77.9 | 22.1 | 1.77 (1.43-2.18)*** | 1.25 (0.95-1.64) | 1.25 (0.92-1.70) |
| Richer | 74.6 | 25.4 | 2.13 (1.73-2.62)*** | 1.34 (1.00-1.78)* | 1.42 (1.02-1.98)* |
| Richest | 67.1 | 32.9 | 3.05 (2.47-3.78)*** | 1.74 (1.29-2.35)*** | 1.66 (1.18-2.34)** |
| **Place of residence** |  |  |  |  |  |
| Rural | 84.3 | 15.7 | 1 | 1 | 1 |
| Urban | 73.4 | 26.6 | 1.95 (1.70-2.23)*** | 1.22 (1.00-1.50) | 0.97 (0.77-1.23) |
| **Antenatal care** |  |  |  |  |  |
| 0 visit | 90.1 | 9.9 | 1 |  | 1 |
| 1 to 3 visit(s) | 83.5 | 16.5 | 1.80 (1.37-2.37)*** |  | 1.64 (1.08-2.47)* |
| 4 visits | 76.7 | 23.3 | 2.77 (2.09-3.67)*** |  | 1.88 (1.23-2.86)** |
| > 4 visits | 68.7 | 31.3 | 4.15 (3.18-5.42)*** |  | 1.89 (1.25-2.86)** |
| **Mode of delivery** |  |  |  |  |  |
| Vaginal delivery | 84.5 | 15.5 | 1 |  | 1 |
| Vaginal delivery assisted by instruments | 83.3 | 16.7 | 1.09 (0.92-1.30) |  | 0.90 (0.74-1.11) |
| Caesarean delivery | 51.7 | 48.3 | 5.10 (4.27-6.08)*** |  | 2.50 (1.89-3.31)*** |
| **Birth attendant** |  |  |  |  |  |
| Doctors and Nurses/Midwives | 66.2 | 33.8 | 1 |  | 0.99 (0.77-1.27) |
| Nobody | 52.5 | 47.5 | 1.77(0.77-4.11) |  |  |
| Doctors | 58.7 | 41.3 | 1.38 (1.13-1.68)** |  | 1 |
| Nurses/Midwives | 86.1 | 13.9 | 0.32 (0.26-0.38)** |  | 0.63 (0.48-0.83)*** |
| Traditional Midwives | 85.9 | 14.1 | 0.32 (0.24-0.44)** |  |  |
| Relatives/Friends/Neighbours | 84.5 | 15.5 | 0.36 (0.25-0.51)** |  |  |
| Another person | 90.1 | 9.9 | 0.22 (0.07-0.71)* |  |  |
| **Postnatal care before discharge** |  |  |  |  |  |
| No | 89.4 | 10.6 | 1 |  | 1 |
| Yes | 71.2 | 28.8 | 3.43 (2.87-4.10)*** |  | 1.15 (0.89-1.49) |
| **Postnatal care within 6 weeks** |  |  |  |  |  |
| No | 90.9 | 9.1 | 1 |  | 1 |
| Yes | 55.2 | 44.8 | 8.07 (6.99-9.32)*** |  | 6.97 (5.89-8.25)*** |
| **Place of delivery** |  |  |  |  |  |
| Home | 14.9 | 85.1 | 1 |  |  |
| Public hospital | 17.6 | 82.4 | 1.22 (0.98-1.52) |  |  |
| Delivery or health centre | 14.2 | 85.8 | 0.95 (0.71-1.26) |  |  |
| Private clinic | 49.2 | 50.8 | 5.53 (4.34-7.04)** |  |  |
| Private surgery | 55.3 | 44.7 | 7.05 (4.10-12.10)** |  |  |
| **Length of stay in a health facility post-delivery** |  |  |  |  |  |
| < a day (hours) | 86.0 | 14.0 | 1 |  |  |
| 1 ≥ days ≤ 7 | 76.3 | 23.7 | 1.92 (1.46-2.51)** |  |  |
| ≥ 1 week | 63.2 | 36.8 | 3.59 (1.93-6.66)** |  |  |

Caption: \*: p<0.05 \*\*: p<0.01 \*\*\*: p<0.001
Model 1: adjusted for sociodemographic and environmental variables
Model 2: adjusted for all variables
LPPC: later postpartum care OR: odds ratios

Age (30–39), higher level of education and higher socioeconomic status were all significantly associated with LPPC (Table 3). Regarding the age factor, women aged 30 to 39 were 23% more likely to use LPPC compared to younger women (aged 15 to 29). Besides, compared to no formal education, primary level and secondary or higher education level increased by 34% and 79% respectively, the likelihood of using LPPC. Therefore, the higher the level of education the higher likelihood of LPPC uptake.

Furthermore, compared to non-attendance to ANC consultations, the likelihood of LPPC uptake increased by 64%, 88%, and 89% for women who received one to three ANC check-ups, four visits, and more than four visits, respectively (Table 3). Thus, the higher the frequency of ANC check-ups, the higher the likelihood of using LPPC.

The type of health professionals who assisted the delivery also had a significant association with LPPC uptake. The presence of only midwives or nurses decreased by 37% the odds of LPPC uptake compared to a delivery assisted by a doctor. Additionally, women who delivered through caesarean were 2.50 times more likely to use LPPC than their counterparts who gave birth through vaginal delivery (Table 3).

Another facilitator of LPPC was the provision of PNC to newborn babies within the six weeks post-delivery. Table 3 indicates that women were almost seven times more likely to use LPPC when their babies received PNC within the same period.

### Factors associated with postpartum morbidity

The findings regarding PPM occurrence are displayed in Table 4.

**Table 4.**
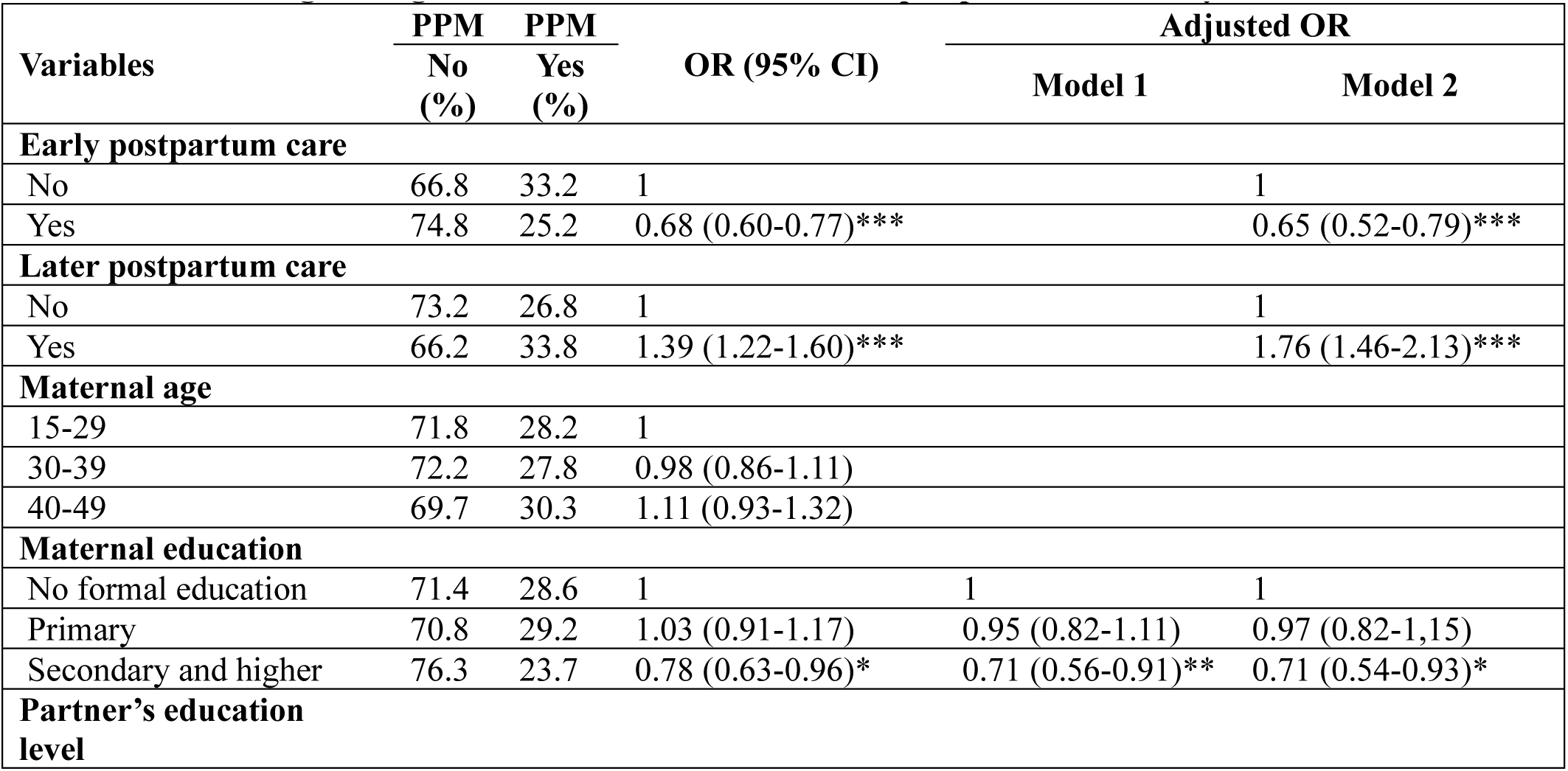
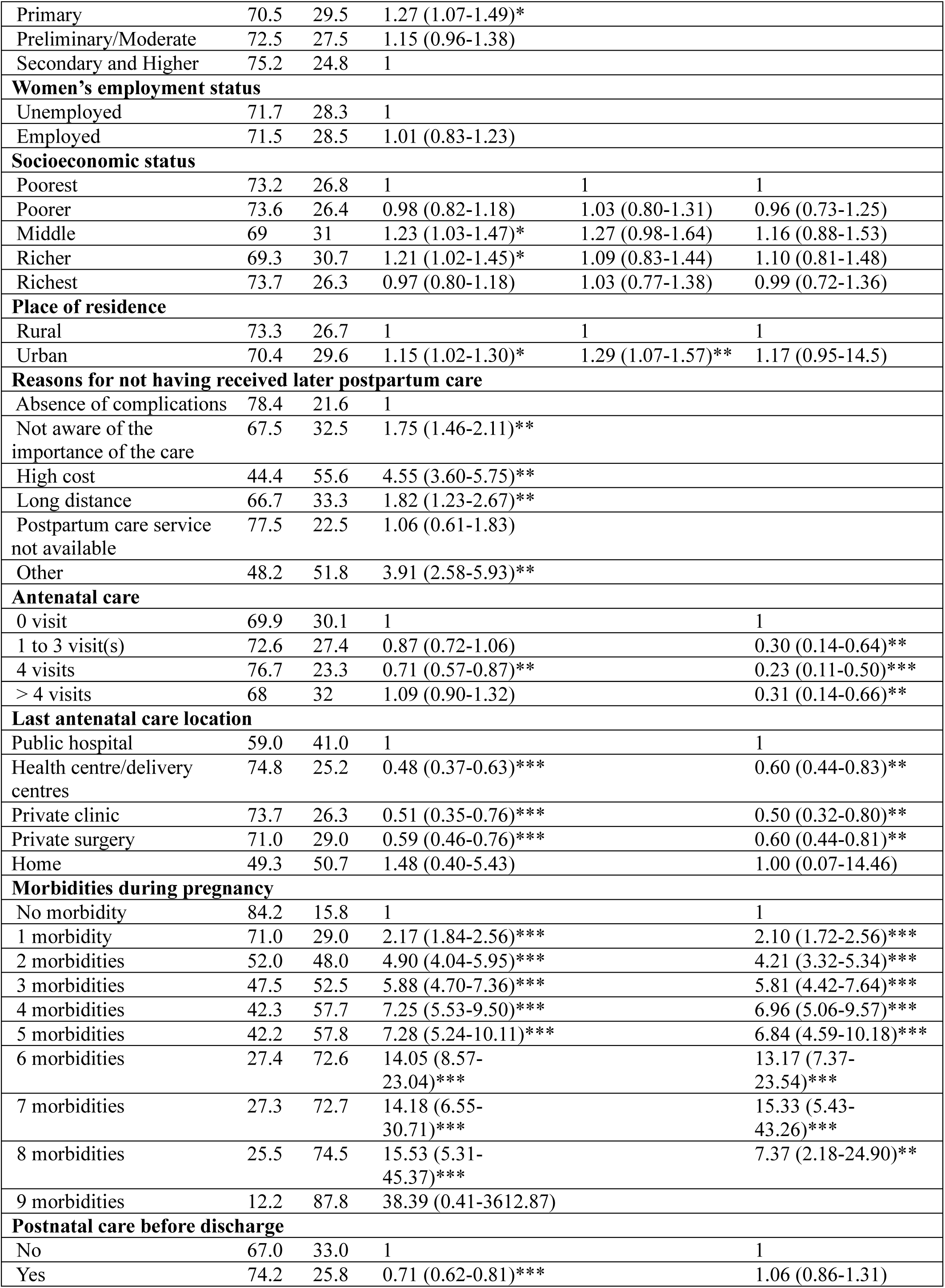
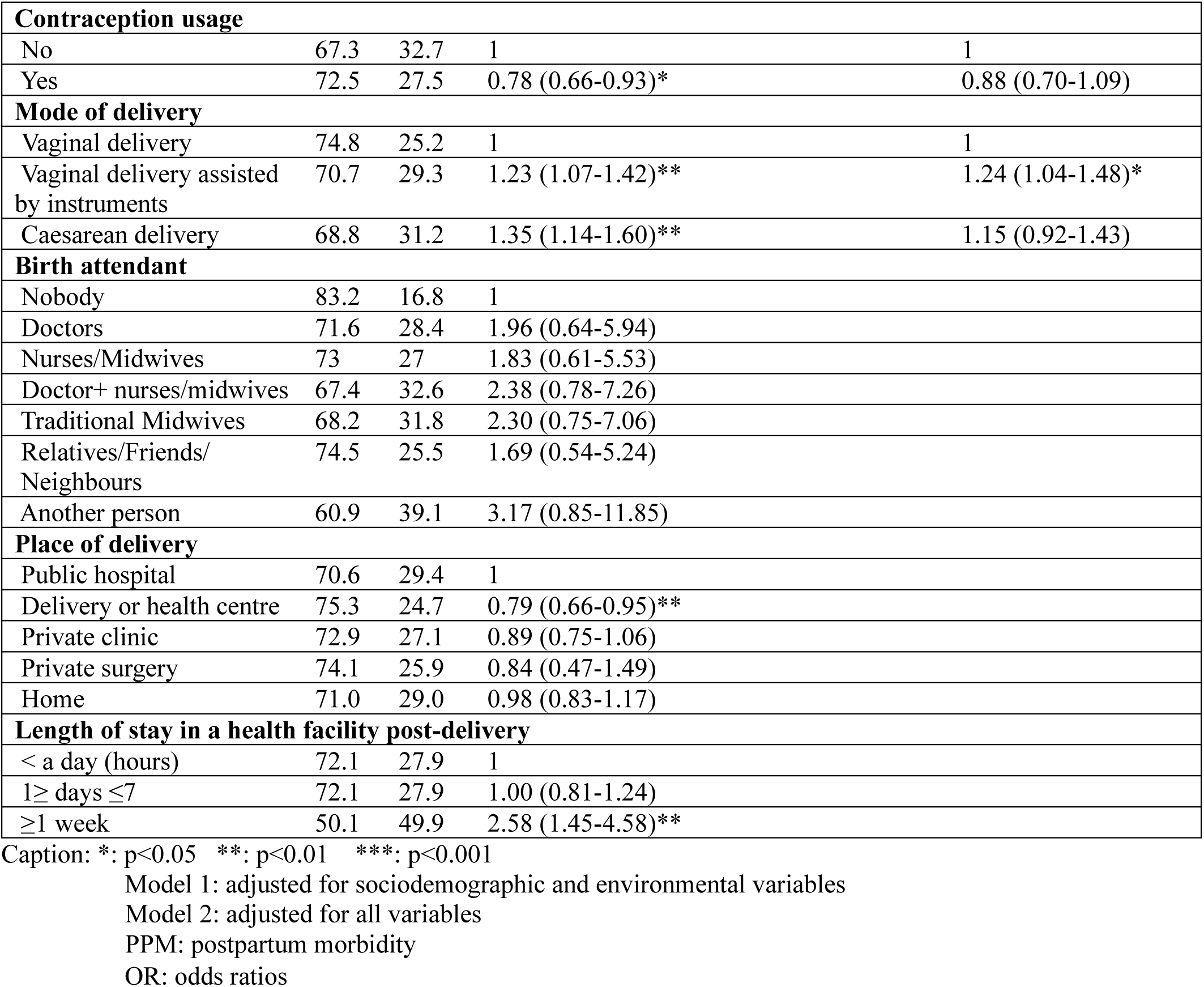
Multilevel logistic regression of factors associated with postpartum morbidity in Morocco.

Among the sociodemographic predictors of PPM, education level was significantly associated with PPM occurrence within six weeks post-delivery. The higher the level of education, the less likely were women to develop PPM. Regarding the obstetric predictors analysed, some appeared to have a protective influence, namely receiving ANC, whilst others were identified as risk factors of PPM, such as the mode of delivery (i.e. instrumental vaginal delivery). Moreover, women who suffered from at least one health issues during pregnancy were significantly more likely to experience PPM. The higher the frequency of pregnancy morbidities, the higher the likelihood of PPM occurrence (Table 4).

### The relationship between EPPC, LPPC and PPM

The relationship between PPC and PPM indicates that receiving EPPC (before discharge) increased by 2.6 times (AOR=2.68, 95%CI:2.08-3.45) the likelihood of LPPC uptake (within 6 weeks) (Table 3) but decreased the risk of PPM occurrence by 35% (AOR= 0.65, 95%CI:0.52-0.79) (Table 4). Thus, EPPC seems to prevent the occurrence of PPM at a later stage. In fact, women who reported one, three, four, and seven PPM were 23% (AOR=0.77, 95%CI:0.61-0.97), 69% (AOR=0.31, 95%CI:0.19-0.50), 77% (AOR=0.23, 95%CI:0.12-0.45), and 96% (AOR=0.04, 95%CI:0.01-0.23) less likely to have received EPPC respectively (Table 2). These observations underline the importance of medical assistance during delivery and the following days.

Furthermore, suffering from one PPM was significantly associated with a 36% (AOR=1.36, 95%CI:1.08-1.71) increased likelihood to receive LPPC (Table 4). Therefore, women seem to consider LPPC check-ups as a curative approach and use it to seek medical assistance if they experience PPM.

## DISCUSSION

### Main findings

The extent of PPC utilisation in Morocco reached 62.6% before discharge and 21.3% within six weeks of delivery. Moreover, 28.3% of PPM (pregnancy-related morbidities) within six weeks post-delivery were reported.

Several sociodemographic factors influenced positively later PPC uptake in Morocco as in other LMIC (10,12), including: being over 30 years old, achieving formal education, and belonging to wealthier households. Women with higher socioeconomic status were more likely to use LPPC, as their financial means enable them to afford continuous monitoring throughout the maternity process in the private sector by the same health professional. This may foster trustful and continuous relationships post-discharge. Education appeared particularly beneficial too, with lower risk of developing PPM and higher odds of LPPC uptake, it contributes to women’s knowledge and awareness of PPC benefits, leading to a proactive health seeking-behaviours (14,15). However, this study (cf. Model 3), did not find significant associations between women’s employment status or residence and LPPC uptake, diverging from findings in others LMIC (16,17).

Among obstetric determinants, the mode of delivery influenced PPC uptake. Caesarean deliveries doubled EPPC and LPPC utilisation, which was similar to the rate reported by other LMIC (16). This may be due to required post-surgical follow-up which might be assimilated to EPPC (18), whereas vaginal delivery, perceived as a natural process, may not prompt the same level of follow-up, especially in heavy workload situations, unless complications arise. Instrumental vaginal delivery was also associated with an increased risk of PPM, consistent with findings from other LMIC reported in the systematic review by Sobhy et al. (19). Procedures such as episiotomy, which is a surgical incision of the perineum to prevent perineal tears often performed during instrumental delivery (20), can increase the risk of puerperal infection; like inappropriate and insufficient self-care practice (21,22). Nevertheless, strengthening training and education of health professionals could enhance the efficiency of resource use and improve women’s postpartum health (23).

The results also showed that PNC provided to newborn babies was the strongest predictor of both EPPC and LPPC. Thus, whether in hospital or within six weeks post-delivery, the care given to babies, such as vaccination, was related to the care provided to women.

The only identified barrier to LPPC uptake was being assisted through delivery by midwives/nurses but without a doctor’s presence. In Morocco, most deliveries occur in public hospitals and are midwife-led for uncomplicated vaginal birth without the assistance of doctors, in line with national regulations (24). However, antenatal and postpartum consultations often delivered by doctors or midwives/nurses are mainly provided in other types of health facilities. In this context, childbirth is an event that generally only involves a short and temporary relationship between women and health professionals. The latter may perceive their responsibilities as limited to care provided within hospital settings, weakening the continuity needed to promote follow-up. Conversely, in private sector, a single health professional often provides care across the maternal health continuum from ANC to LPPC. Their duty of care focuses on the woman rather than the facility, which encourage them promoting LPPC consultations. Besides, giving birth in a health facility, in particular private structures, also appeared as a potential facilitator of EPPC and LPPC uptake (cf. univariate analysis), a trend also observed in Zambia (25). These results emphasize the importance of the health professional-woman relationship in ensuring continuity of care (26).

Furthermore, the results indicated that attending four ANC consultations was positively associated with EPPC and LPPC uptake, and reduced PPM risk, underscoring the important role of antenatal counselling in promoting LPPC to manage postpartum issues. However, women who experienced health issues during pregnancy were at greater risk of developing PPM. This finding corroborates previous studies on maternal near-miss cases (27,28), which refer to instances where women survive severe, life-threatening complications during pregnancy, childbirth, or within six weeks post-delivery (29). Ensuring effective pregnancy monitoring can therefore improve PPC uptake and help reduce maternal morbidity and mortality (30).

### What is already known on this topic

PPC is among the most neglected components of maternal healthcare in LMICs, despite its importance in preventing and managing PPM. Existing studies have identified key sociodemographic and obstetric facilitators of PPC such as wealth, education, urban residence, and caesarean delivery. Moreover, instrumental deliveries can increase the risk of complications. However, gaps in continuity between delivery and postpartum care, particularly in public facilities, are common. The relationship between health professionals and women, and the integration of maternal and newborn services, have also been recognised as important in encouraging PPC follow-up.

### What this study adds

The novel contribution of this study lies in its use of nationally representative data from Morocco to generate new insights into PPC uptake, which may also be relevant to other LMIC. The study confirms established facilitators of PPC (e.g., education, ANC visits), but also highlights system-level factors affecting care continuity. Specifically, the absence of a doctor during delivery was linked to reduced LPPC attendance, suggesting that midwife or nurse-led care in public hospitals may lack the follow-up mechanisms necessary for encouraging PPC visits.

The strong association between newborn care and maternal PPC use suggests that integrating mother– child services may be an effective strategy to boost PPC. Another key aspect explored is the temporal dynamics of care: EPPC prevents later PPM and promotes LPPC, while LPPC is often used by women as a recourse in response to postpartum health symptoms. These findings have practical implications for improving maternal health programs in Morocco and similar contexts.

### Limitation of this study

This study presents several limitations. Retrospective data collection may have introduced recall bias, while the exclusion of cases with missing data may have resulted in selection bias. Moreover, data were self-reported without medical verification to corroborate women’s declarations, possibly leading to underreporting, particularly of psychological morbidities which were not addressed in the questionnaire. Similarly, other important aspects including cultural practices, beliefs, and women’s autonomy in decision-making were not cover in the survey which may have limited the depth of insight. Furthermore, data lacked clarity about the number of PPC check-ups, hindering any comparison with the WHO guidelines (i.e. four PPC check-ups within six weeks). Lastly, findings must be interpreted cautiously because causality relationships cannot be concluded due to the cross-sectional study design.

## CONCLUSION

PPC utilisation remains low in Morocco, particularly later PPC (21.8%) while PPM persist. Both variables were marked by social health inequities with a clear social gradient based on education and socio-economic status. This study showed that pregnancy monitoring encouraged continuity of care and reduced PPM occurrence. Ultimately, the findings support the need of efficient interventions promoting PPC uptake. A qualitative study may help to gather insights on the reasons that encourage women to use later PPC and their perception of postpartum care and morbidities.

## Data Availability

All data produced in the present work are contained in the manuscript.

## Acknowledgement

We thank the Division of Planning and Study of the Moroccan Ministry of Health for sharing the data analysed in this study.

This study is part of the first author’s PhD thesis at the University of West London. The thesis is available at the university repository:

(https://repository.uwl.ac.uk/id/eprint/10597/1/Habib%20%20Final%20PhD%20Thesis%20(Dec%2023).pdf).

