## Supplementary figures and images for "The relationship between postpartum care uptake and postpartum morbidity and their determinants in Morocco: evidence from a national survey"

### Supplementary file 1

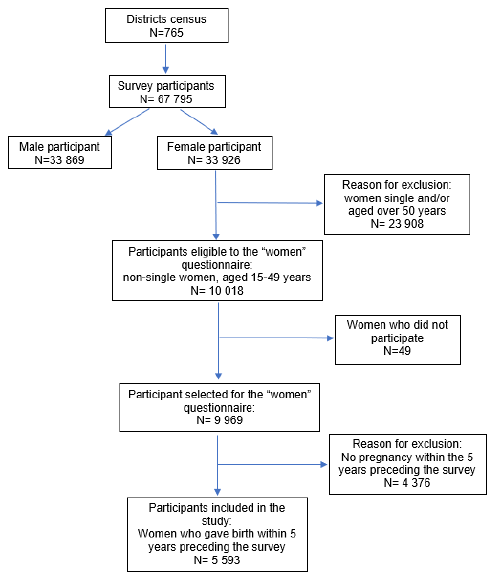

**Supplementary file 1. Flowchart of the selection process of the study participants**
